# A Systematic Review of the Incubation Period of SARS-CoV-2: The Effects of Age, Biological Sex, and Location on Incubation Period

**DOI:** 10.1101/2020.12.23.20248790

**Authors:** Caitlin Daley, Megan Fydenkevez, Shari Ackerman-Morris

## Abstract

A systematic review of the incubation period of COVID-19 was compiled and analyzed from 21 quantitative studies. We investigated the incubation period of COVID-19 with regard to age, biological sex, location, and severity of the disease. Based on the data extracted, we report an overall mean and median incubation period for SARS-CoV-2 of 5.894 days and 5.598 days, respectively. The incubation period did not statistically vary for biological sex or age, but some studies suggest a longer incubation period in the young and elderly. Cases of COVID-19 in Wuhan and Hubei Province of China may have a shorter incubation period for COVID-19 but the shorter incubation period may be due to an increase in viral load. In studying coronavirus strains such as SARS and MERS, researchers have discovered an inverse relationship between incubation period length and virus severity. Taking into consideration that SARS-CoV-2 is part of the beta-coronavirus family, as well as the study mentioned above, we suggest that people who experience more severe disease due to SARS-CoV-2 may have a shorter incubation period.

## INTRODUCTION

In late December of 2019, several patients presented with pneumonia of an unknown cause in Wuhan City of the Hubei province in China (28). Using throat swab specimens, the causative agent of these cases was soon identified as a novel coronavirus, which was later named by the World Health Organization (WHO) as COVID-19, and by the International Committee on Taxonomy of Viruses (ICTV) as SARS-CoV-2 (28, 10). Following sequencing, SARS-CoV-2 was classified as a beta-coronavirus, a subclass of the Coronaviridae family (10, 28). Further sequencing analysis, as well as phylogenetic analysis, has demonstrated a high degree of similarity in sequence between SARS-CoV-2 and SARS-CoV and MERS-CoV, which are also classified as beta-coronaviruses (10, 26). While there are many notable similarities in clinical presentation among these three classes of coronaviruses including fever, dry cough, and dyspnea, SARS-CoV-2 has been linked to a unique attack of the lower airway, causing additional symptoms such as rhinorrhea, sneezing, and sore throat (26).

Primarily targeting the respiratory system, SARS-CoV-2 hijacks the human angiotensin-converting enzyme 2 (ACE2) in the epithelial cells of the lungs, which normally functions by cleaving angiotensin I to produce angiotensin II (10, 26). Acting as a binding site for the spike proteins of coronaviruses, ACE2 binds to the SARS-CoV-2 spike protein with an affinity 10–20 times higher than that of SARS-CoV, potentially making SARS-CoV-2 more virulent and dangerous to the population (10).

Although the origin of the virus has been linked to Wuhan’s Huanan Seafood Wholesale Market, with bats acting as an alternative host, the disease has quickly spread out of China, resulting in a global pandemic (27, 28, 26, 10). As of July 31, 2020, the WHO reported 17,064,064 confirmed cases of COVID-19, including 668,073 deaths, globally (35). Even with vaccines starting to be available, prevention is essential. Prevention relies on understanding the transmissibility of the virus, specifically the incubation period.

Incubation period is defined as the time between exposure to the pathogen and the onset of signs or symptoms. The length of this period is determined by several properties: the amount of infectious viral particles one is exposed to, the rate of viral clearance by the host’s innate and adaptive immune systems, and the impact of viral evasion and the viral load (12). Incubation period is critical, as it can help dictate how well practices such as contact tracing and entry screening are functioning (2).

Furthermore, the incubation period is an important component of containing the transmission of COVID-19. Significant concerns have lingered throughout the world about when to re-open work places, schools, and restaurants, as well as when to hold large public gatherings such as concerts, weddings, and festivals. The re-opening of these features of society all partly rely on a set quarantine period (6). For SARS-CoV-2 the quarantine period has been set for 14 days. This is based on research that shows the median incubation period to be 5.1 days and 97.5% of patients who developed symptoms did so within 11.5 days and 99% developed symptoms within 14 days (16). As the incubation period of a virus has the potential to differ in length, this is vital information in determining if the current quarantine period should be adjusted after possible exposure to the viral pathogen. For an appropriate length of quarantine period to be determined, an accurate, uniform incubation period should be established (6).

During the incubation period, individuals that are infected still have the ability to be contagious, and therefore transmit the virus without knowing they have been infected. This period is also known as the pre-symptomatic stage. Transmission during this stage has been documented through contact tracing efforts. To support the existence of the pre-symptomatic stage of contraction, data has shown that some can test positive between 1-3 days after contraction (34).

Another key component of containing the virus is the recognition of asymptomatic transmission and contraction. Asymptomatic cases are those that have been laboratory tested positive for SARS-CoV-2 but have not developed symptoms. Asymptomatic cases can still transmit disease during the early days of transmission like non-asymptomatic cases. With an accurate, uniform incubation period established, the transmission of asymptomatic cases could be lowered (34).

Gaining a comprehensive understanding of a disease is contingent on identifying the vulnerable populations. In the era of quarantining and social distancing, it may become critical to tailor preventive measures according to at-risk groups as the world begins to resume a “new-normal” function. This includes analyzing the incubation period of distinct population groups. While it is known that specific age groups, specifically the elderly population, may be at a greater risk for infection from SARS-CoV-2, differences in incubation period among age groups remains a topic of discussion. An age-stratified understanding of incubation period may permit more specific quarantine guidelines and preventative measures, particularly for the schools. Additionally, it is apparent that certain medical conditions disproportionately affect the biological sexes. In terms of SARS-CoV-2, analyzing differences in the incubation period between sexes contributes to a more concise understanding of the transmissibility of the disease.

An inverse relationship between incubation period length and severity of disease has been found related to other beta-coronaviruses including MERS-CoV and SARS-CoV. In a study on the association between severity of MERS-CoV infection and incubation period, researchers found patients who died from the disease caused by MERS-CoV had a shorter incubation period while patients who lived had longer incubation periods. In a previous study by the same researchers, the same correlation between severity and incubation period was found related to the SARS-CoV virus (31). Correlation between severity and the incubation period of SARS-CoV-2 is currently still under investigation.

Since the outbreak of the virus in Wuhan, researchers around the globe have set out to determine the incubation period of the novel coronavirus. Due to the novelty of the virus and the subsequent lack of information regarding its clinical characteristics and transmission dynamics, researchers have been compiling case information as it becomes available, most often using government case reports. In this systematic review, research regarding the incubation period of COVID-19 was compiled and analyzed using conventional and artificial intelligence (AI)-assisted search strategies with the goal of investigating the links among incubation period and age, biological sex, location, and severity of the disease.

## METHODS

### Databases and Search Strategies

Between the dates of June 16th and July 18th 2020, databases including PubMed, Science Direct, and MedRxiv, were searched for primary research articles pertaining to the incubation period of SARS-CoV-2 that had been published since the outbreak of the virus. Different combinations and variations of search terms were employed in order to best represent any articles investigating the clinical characteristics of the virus (e.g. “COVID-19”, “COVID19”, “novel coronavirus”, “SARS-CoV-2”, “incubation period”, “clinical features”, “transmission dynamics”, “severity”, “location”, “age”, “gender”, “biological sex”). In addition to these search strategies, a natural language AI-powered search engine created by Dr. Tayab Waseem at Eastern Virginia Medical College was used to generate a list of all published SARS-CoV-2 literature that answered the research question, “What is the incubation period of SARS-CoV-2?”. The AI search engine searched all scientific research articles available online up to June 15, 2020.”

### Inclusion Criteria

Quantitative studies were included if they met the following inclusion criteria: defined the incubation period as the time between exposure to the virus and onset of symptoms, employed contact tracing to identify incubation period timeline, only included cases with precise dates of exposure and symptom onset, confirmed potential cases using reverse-transcriptase polymerase chain reaction (RT-PCR) with nasal/throat swabs or blood specimens, and only included confirmed SARS-CoV-2 cases. Studies that did not meet these criteria were excluded from analysis.

### Data Extraction

For each study included in the analysis, study type, sample size, age and sex of patients, location, incubation period statistics, and citation information were recorded.

### Data Analysis

#### Overall Incubation Period

Using the data from the 21 compiled quantitative studies, the studies that reported a mean incubation period were used to estimate an overall mean incubation period for SARS-CoV-2. This was done by taking an average of the mean incubation periods reported for a total of 8 studies that reported their incubation period as such. A similar procedure was used to generate an overall estimation of the median incubation period; an average of the median incubation periods of 8 studies that reported their incubation period as a median was taken.

#### Age

Using the studies that reported their incubation period as a mean, the median age of every age group studied, for each study analyzed, was taken in order to generate one plottable point corresponding to a specific age. This median age was plotted against the corresponding reported mean incubation period for that age group to generate one scatter plot depicting the relationship between age and incubation period for all studies analyzed (see Figure 1). The same procedure was used for those studies that reported their incubation period as a median, as seen in Figure 2.

**Figure 1:**
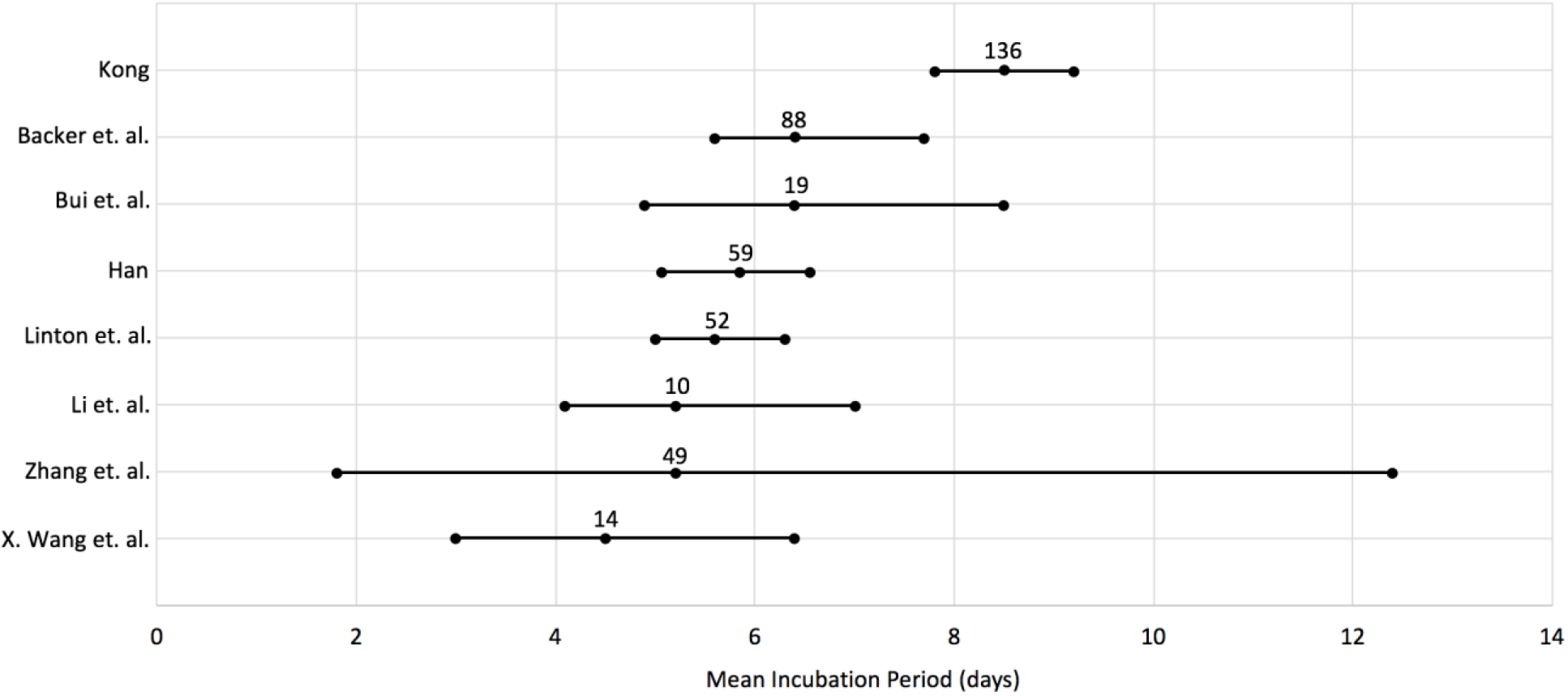
Forest plot of the mean incubation period and 95% confidence interval for 8 quantitative studies that reported their incubation period in this manner. The number above the central point for each study represents the sample size of that study.

**Figure 2:**
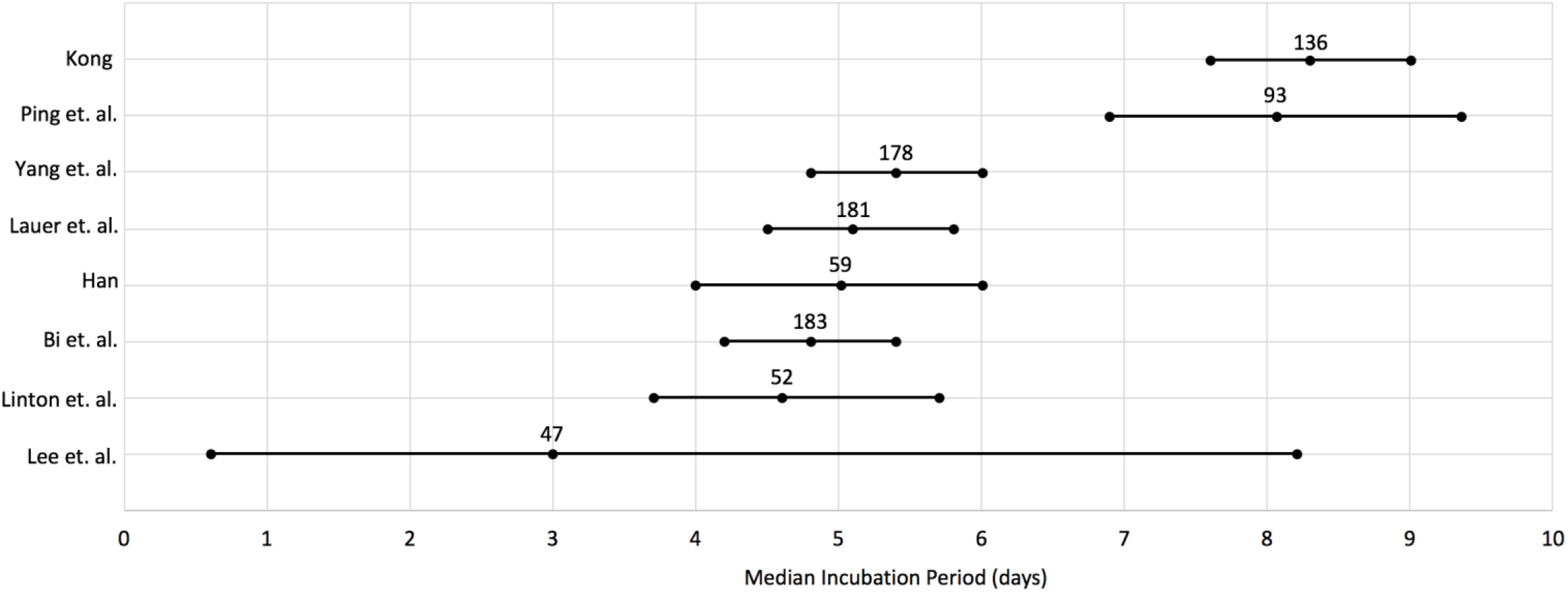
Forest plot of the median incubation period and 95% confidence interval for 8 quantitative studies that reported their incubation period in this manner. The number above the central point for each study represents the sample size of that study,

## RESULTS

### Incubation Period

Database searches generated 21 primary studies that fit the inclusion criteria and were included in the analysis of overall incubation period, as shown in Table 1. Based on the data from these quantitative studies, we report an overall mean incubation period of 5.894 days and an overall median incubation period of 5.598 days. Forest plots of those studies that reported the incubation period as a mean with a 95% confidence interval or a median with a 95% confidence interval, are shown in Figures 1 and 2 respectively.

**Table 1:**
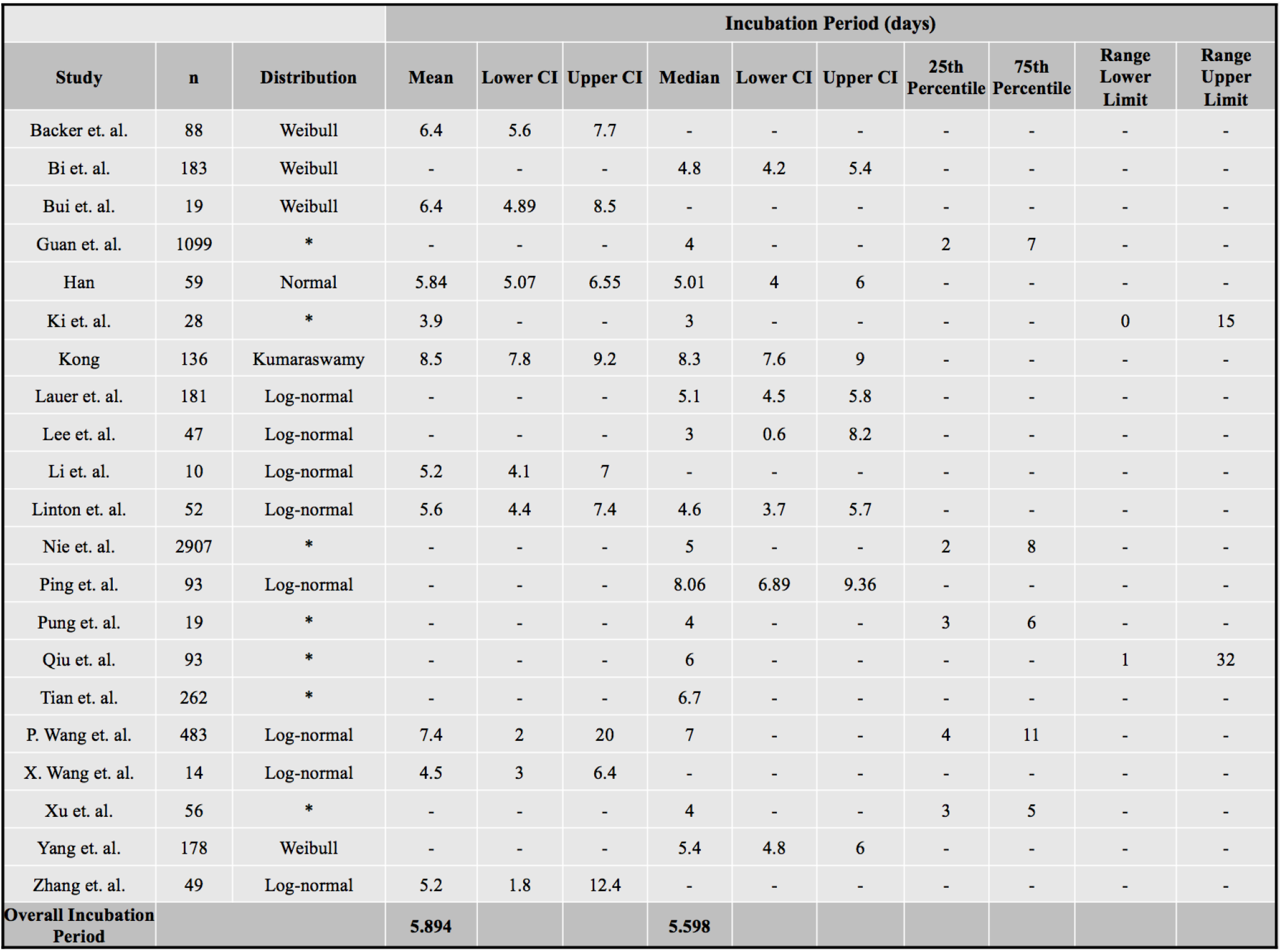
Compilation of 21 studies used for the analysis of overall incubation period of SARS-CoV-2. Studies with an asterisk in the distribution column indicate the use of descriptive statistics only.

### Age and Incubation Period

Several studies have investigated the incubation period of SARS-Cov-2 according to age groups. In a study conducted on pediatric patients in China, 12 patients under the age of 18, ranging in age from 7 months to 17 years, were recruited from several local hospitals in the Chongqing area. Results revealed an average incubation period of 8 days, which was found to be considerably longer when compared to a study of adult patients who exhibited an average incubation period of 3 days (5). Additionally, a study conducted by Zhang et.al. investigated the incubation period of 49 patients aged 33-56, otherwise classified as young to middle aged adults. Results revealed a mean incubation period of 5.2 days (40).

Additional studies have divided the population into specific age groups for further analysis. One particular study found age 40 to be a key cutoff point in incubation period. The median incubation period of patients under the age of 40 was found to be significantly different than that of patients ages 40 years and older, with the older age group exhibiting a longer median incubation period. Furthermore, these age groups were found to be linearly separable, which did not hold true when the age groups were divided as ages 50 and older and less than 50, or as ages 55 and older and less than 55 (8). An additional study investigated the incubation period among adolescents and young adults. Defining adolescents as 10-24 years of age and young adults as 25-35 years of age, researchers analyzed the medical records of 46 patients within the age constraints. A median incubation period of 6.6 days was found across both age groups (18).

Yet, several other studies have further divided the population into additional age groups, presenting a more detailed and comprehensive breakdown. One study conducted at the epicenter of the pandemic investigated the differences in incubation period across a range of ages, divided into age groups with a size of 15 years. Results revealed that incubation period exhibits a U-shaped curve relative to age group, with pediatric and geriatric patients experiencing a longer median incubation period. More specifically, younger adults, designated as ages 15-64, demonstrated a shorter median incubation period (7.6 days) than older adults (11.2 days), which were designated as ages ≥65 (14). In a similar study, incubation period was investigated across 4 different age groups: <14, 14-34, 35-64, and >64. While no significant difference was found between age groups, the data exhibited a U-shaped curve with patients on either extreme experiencing longer median incubation periods, as demonstrated similarly by the study conducted by Kong (39). The incubation period reported by each study, as well the incubation periods reported for each individual age group studied, is shown in Table 2.

**Table 2:** Incubation period broken out by age.

|  |  |  | Incubation Period |  |
| --- | --- | --- | --- | --- |
| Study | n | Age | Median | Mean |
| Chen et. al. | 12 | 7 months - 17 years | - | 8 |
| Han | 59 | <40 | 4 | 4.84 |
|  |  | ≥40 | 6 | 6.73 |
| Kong | 136 | 0-14 | 13.5 | 12 |
|  |  | 15-29 | 9.5 | 8.9 |
|  |  | 30-44 | 8.5 | 8.4 |
|  |  | 45-59 | 6 | 7.2 |
|  |  | 60-74 | 9 | 9.6 |
|  |  | 75-89 | 13 | 13.2 |
|  |  | younger adults: 15-64 | 7.6 | - |
|  |  | older adults: ≥ 65 | 11.2 | - |
| Liao et. al. | 14 | 10-35 | 6.6 | - |
| Yang et. al. | 178 | <14 | 8 | - |
|  |  | 14-34 | 5 | - |
|  |  | 35-64 | 6 | - |
|  |  | >64 | 9 | - |
| Zhang et. al. | 49 | 33-56 | - | 5.2 |

Using the median age of each age group studied, and the corresponding median incubation period, the data from four of the six studies were plotted together as a scatter plot, as shown in Figure 3. The two studies that did not report their incubation period as a median were excluded from this plot. The data from the Kong study exhibits a strong U-shaped curve, with the data from the Yang et. al. study exhibiting a slightly weaker and less defined version of this pattern. When viewed all together, the data does not exhibit a strong U-shaped curve, suggesting that more research must be conducted to confirm this pattern.

**Figure 3:**
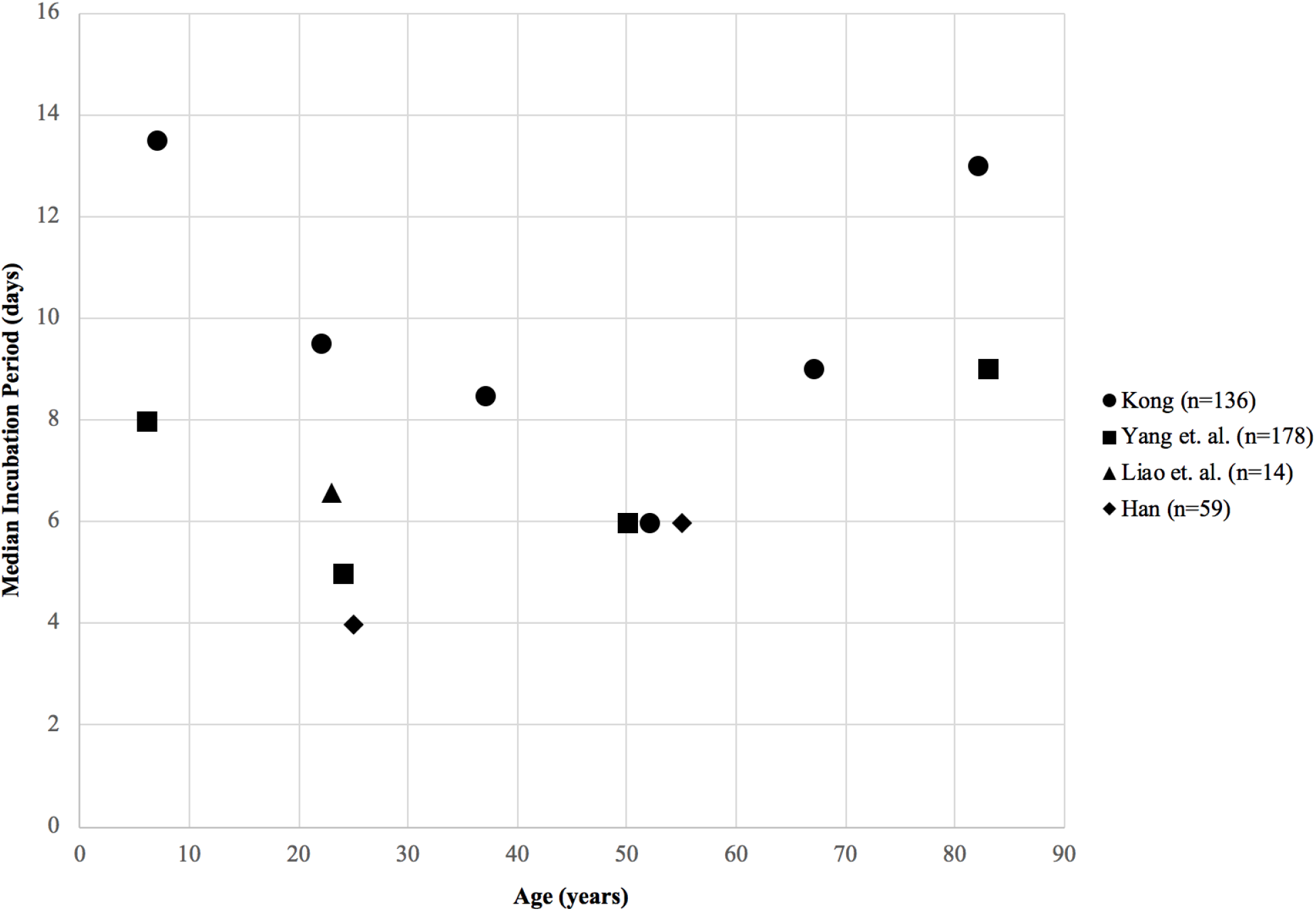
Scatter plot showing the median incubation period of SARS-CoV-2 relative to the median age within each age range studied for four different studies. Individual studies are distinguished by shape, as shown in the legend. It is important to note a weak U-shaped curve formed when taking all the data points into account, with the data from Kong and Yang et. al. constituting the backbone of the curve.

### Biological Sex and Incubation Period

Several studies that have investigated the incubation period of SARS-CoV-2 have also analyzed the relationship between biological sex and the incubation period. A study conducted in China found that males have a slightly shorter median incubation period compared to females, however this difference was not determined to be statistically significant (8). Another study found similar results, with males exhibiting a slightly shorter, but not significantly different, median incubation period than females (16). Contrastingly, a study conducted by Nie et. al. found just the opposite: that men have a slightly longer incubation period than females, however this difference was not found to be statistically significant (22). Yet, another study found no differences at all in the incubation period of males and females (39). The incubation period reported by each study, including the individual incubation periods reported for males and females, is shown in Table 3.

**Table 3:** Individual incubation periods reported for males and females

|  | **Median Incubation Period** |  | **Statistical Significance** |
| --- | --- | --- | --- |
| **Study** | **Male** | **Female** | **p-value** |
| Han | 5 | 5.5 | 0.267 |
| Nie et. al | 5 | 4 | 0.22 |
| Lee et. al | 2.5 | 3.3 | 0.3 |
| Yang et. al | 6 | 6 | 0.848 |

### The Effects of Travel on Incubation Period within China

In a study on the clinical features and dynamics of viral load in imported and non-imported patients with COVID-19, researchers found that people who had not visited Wuhan or had come into contact with people from Wuhan, also known as the tertiary group, had a longer incubation period than both the secondary and imported groups (p < 0.01 and p = 0.03, respectively). The imported group included people who had acquired COVID-19 that had visited or originated from the city of Wuhan, while the secondary group did not originate from or visit Wuhan but rather came in contact with imported cases (37).

In Char Leung’s study from Deakin University, differences in the incubation period of COVID-19 between people who traveled to Hubei (Province where Wuhan is located) and those who did not were examined. They found a significant difference in the distribution of the incubation period between travelers to Hubei and those who did not travel to the province. The mean incubation period for non-travelers was reported at 7.2 days, while the traveler’s mean incubation period was reported at 1.8 days. The suggested reasons for the difference between the two groups are location and variability; variance was noted to possibly be linked to the infectious dose for the travelers to Hubei, and the group of travelers was said to have had multiple different sources of infection while visiting Hubei. The higher variance in incubation periods may result in a longer recommended quarantine period (18).

Similarly, Zuopeng Xiao’s study from the Harbin Institute of Technology examined the incubation period of COVID-19, including people infected within Wuhan, relative to local transmission outside of Wuhan. Local transmission outside of Wuhan exhibited longer incubation periods than “early generation patients” who were directly exposed to Wuhan. It was found that patients who regularly lived in Wuhan and left to other locations before January, had a shorter posterior median value for incubation period (7.57 days), whereas the group that did not travel and was affected by local transmission outside of Wuhan had a longer posterior median value of 9.31 days (36). Lastly, in a study conducted in Guangzhou, China, it was found that there was no difference between patients who likely contracted the virus in Wuhan versus outside of Wuhan (33).

## DISCUSSION

At the present time, numerous studies have been conducted regarding the incubation period of SARS-CoV-2. Based on the data extracted from 21 quantitative studies, we report an overall mean and median incubation period of 5.894 days and 5.598 days, respectively, for this virus. These findings are consistent with the findings of several meta-analyses that analyzed the overall incubation period of SARS-CoV-2; Kahlili et. al. reported a mean incubation period of 5.84 days, while McAloon et. al. reported a mean incubation period of 5.48 days (12, 21).

In terms of age breakdown, the data from Kong and Yang et. al. specifically demonstrate that the incubation period of SARS-CoV-2 exhibits a U-shaped curve, with Kong exhibiting a slightly stronger U-shaped pattern. According to this pattern, pediatric and geriatric populations may experience longer incubation periods than those ages that fall between these divisions (14, 39). Furthermore, Han found a significant difference in incubation period in patients less than 40 years of age and ages 40 and older (8).

However, each study utilized different age boundaries, and included a different number of ages within these boundaries. While Kong (14) consistently divided age groups by 15-year intervals, Han (8) had an essentially equal split into 2 age groups, and Liao et. al. (18) studied one set age group, Yang et. al. (39) created age groups of notably different sizes. Furthermore, Yang did not note the full range of ages studied, thus ages “<14” and “>64” do not produce definitive boundaries (39). Future research investigating age and incubation period should standardize age divisions, especially age group definitions such as “pediatric,” “adolescent,” “young adult,” and “geriatric.” When examining the compilation of data from Kong (14), Yang et. al. (39), Liao et. al. (18), and Han (8), as shown in Figure 3, incubation period appears to exhibit a weak U-shaped curve relative to age, however it is important to note the data from Kong and Yang et. al. makes up the majority of this curve. Additionally, the 20-30 years age range exhibited the most inconsistencies in the incubation period. This may suggest a high degree of variability in the incubation period within this age group. At this time, more research is needed in order to determine any significant trends regarding the relationship between age and incubation period.

Longer incubation periods in specific age populations may warrant additional social distancing measures for these groups in order to limit the spread of the virus throughout the entirety of the population. As schools reopen for in person instruction, understanding the incubation period of these respective age groups must be a critical piece of the plan. Elementary and middle schools may require additional precautionary measures, as their students fall within an age range that experiences a longer incubation period, which may permit more spread if the individual is unaware, they are infected for a longer period of time. This age group may also require a longer quarantine period. On the other hand, colleges and universities may have comparatively less of a concern, as these students have a shorter incubation period, less continuous contact with peers, and fewer shared classroom materials. However, administrators and professors may complicate planning at the collegiate level, as they may exhibit a longer incubation period than students due to their age. Concerning the geriatric population and nursing homes, without routine testing, individuals may not know they are infected for a long period of time due to the characteristically longer incubation period of this age group. As this population is considered to be at high-risk for SARS-CoV-2, delayed identification can be fatal. This may warrant prolonged implementation of safety measures, including limited contact with members of the nursing home, as well as limited contact with the outside community.

Although some studies have found slight differences in the incubation period of males and females, these differences have not been found to be significant (8, 16, 22). Thus, at this time, biological sex does not appear to play a role in the incubation period of SARS-CoV-2.

The majority of the research that has been conducted specifically on locations related to the incubation period of SARS-CoV-2 has focused on the province of Hubei, specifically the city of Wuhan. In comparing the studies identified in this paper, the findings suggest that in Wuhan, the origin city of the virus, individuals have experienced shorter incubation periods than those outside of Wuhan. Similarly, in the province of Hubei patients have experienced shorter incubation periods than outside of that specific province. However, research by Wang and associates stated that they found no difference in incubation period between patients who contracted the virus in Wuhan versus outside of Wuhan (33). Based on these different findings regarding the incubation periods in the Hubei province and city of Wuhan, further research will be required to determine the causality of these effects.

Through previous research of other coronavirus strains such as SARS and MERS, researchers have discovered an inverse relationship between incubation period length and virus severity (11). Analyzing data from 2003 from Hong Kong has shown that a shorter incubation period has been associated with more-fatal cases of the virus (11). A similar outcome was found in 2015 during the MERS-CoV outbreak in South Korea. Patients who had died from disease caused by the MERS virus experienced a shorter incubation period (11). At this time there is limited published literature on incubation period related to severity of COVID-19, however one study by Yaping Wang and a team from Guangzhou Medical University researched the clinical characteristics of patients infected with SARS-CoV-2 in Guangzhou, China. Authors of this study found that patients with severe disease had a shorter incubation period (33). Taking into consideration that SARS-CoV-2 is part of the beta-coronavirus family, as well as the study mentioned above, we suggest that people who experience more severe disease due to SARS-CoV-2 may have a shorter incubation period.

Amidst the continual spread of the global COVID-19 pandemic, countries have begun to reopen their economies and relax social-distancing guidelines. It is therefore essential to understand the incubation period of the virus for the construction of evolving quarantine guidelines and the prevention of future spikes in infection rates.

## Data Availability

All data used in this study are appropriately references.

## Acknowledgements

We would like to thank the following people for their help. Augustina Nguyen for research, summaries and editing on the paper. Helder Prece, Sara Al Yasin, Madeline Michard, Claire Avril, Rebeca Houghton and Sydney Sipes for research and paper summaries and Anoushka Agrawal and Chole Deubner for research. We would also like to thank Dr. Kathleen Morgan and Dr. Hilary Gaudet for their invaluable input on this process and Jillian Amaral for showing us the best practices for finding relevant research papers. A big thanks to Dr. Tayab Waseem for the artificial intelligence search results and Dr. Robert Morris for leading this project at Wheaton College, MA.

